# Pregnancy, fertility concerns, and fertility preservation procedures in French breast cancer survivors in the FEERIC national study (on behalf of the Seintinelles research network)

**DOI:** 10.1101/2021.06.07.21258472

**Authors:** Manon Mangiardi-Veltin, Clara Sebbag, Christine Rousset-Jablonski, Isabelle Ray-Coquard, Clémentine Berkach, Lucie Laot, Yuting Wang, Inès Abdennebi, Julie Labrosse, Camille Sautter, Aullène Toussaint, Laura Sablone, Enora Laas, Sarah Khallouch, Florence Coussy, Pietro Santulli, Charles Chapron, Angelique Bobrie, William Jacot, Nadir Sella, Elise Dumas, Claire Sénéchal-Davin, Marc Espie, Sylvie Giacchetti, Lorraine Maitrot, Geneviève Plu-Bureau, Charles Coutant, Julien Guerin, Bernard Asselain, Pierre Fumoleau, Manuel Rodrigues, Christine Decanter, Audrey Mailliez, Lidia Delrieu, Seintinelles research network, Fabien Reyal, Anne-Sophie Hamy

## Abstract

**Objective:** To study fertility concerns and oncofertility practices at time of breast cancer (BC) diagnosis.

**Design:** The FEERIC study (Fertility, Pregnancy, Contraception after BC in France) is a prospective, multicenter study.

**Setting:** Web-based collaborative research platform *Seintinelles*.

**Patients:** 517 patients with prior BC diagnosis free from relapse and aged 18 to 43 years at inclusion (from 12^th^ March 2018 to 27^th^ June 2019).

**Intervention:** Baseline online self-administered questionnaires.

**Main Outcome Measure:** Fertility preservation procedures at BC diagnosis.

**Results:** Median age at BC diagnosis was 33.6 years and 424 patients (82.0%) received chemotherapy. Overall, 236 (45.6%) patients were offered specialized oncofertility counseling, 124 (24.0%) underwent one or more FP procedures with material preservation (oocytes n=108, 20.9%; embryos n=31, 6.0%; both oocytes and embryos n=13, 2.5%; ovarian cryopreservation n=6, 1.2%) and 78 patients received gonadotropin-releasing hormone agonists (15.1%). With a median follow-up of 26.7 months after the end of treatments,133 pregnancies (25.7%) had occurred in 85 patients (16.4%), including 20 unplanned pregnancies (15.0%). Most of the pregnancies were spontaneous (n=113, 87.6%), while 16 (12.4%) required medical interventions. Patients who had an unplanned pregnancy were less likely to have received fertility counseling (p=0.02) and contraceptive counseling (p=0.08) at BC diagnosis.

**Conclusion:** Most of the patients were not offered proper specialized oncofertility counseling at the time of BC diagnosis. Spontaneous pregnancies after BC were very much more frequent than pregnancies resulting from the use of cryopreserved gametes. Adequate contraceptive counseling seems as important as information about fertility and might prevent unplanned pregnancies.

## Introduction

Breast cancer (BC) is the most common **cancer among young women** (1), with approximately 6000 women under the age of 45 years diagnosed in France every year (2). Primary BC treatments include surgery, chemotherapy, radiotherapy and targeted therapy as hormonal therapy and trastuzumab. BC treatments can have a direct (chemotherapy-induced ovarian damage) or indirect (fertility decline due to delayed pregnancy project) impact on fertility (3). In the general context of a trend towards women delaying their first pregnancies in the Western world, increasing numbers of women are being diagnosed with BC before they have had their children (4). Furthermore, BC survival rates are increasing (2), and survivorship issues (including motherhood after cancer) are increasingly being encountered in routine practice. In addition, recent studies have confirmed the long-term safety of pregnancy after BC independently of hormonal receptor status (5,6), *HER2* status (7) or the presence of a *BRCA* mutation (8).

**Fertility preservation (FP) procedures** have developed rapidly over the last decade and are now part of the supportive care offered to patients in routine practice. Available FP methods (9) include (i) the retrieval and storage of frozen material sampled before the start of BC treatment (mature oocyte or embryo cryopreservation, ovarian-tissue cryopreservation, and immature oocyte cryopreservation after *in vitro* maturation (IVM)); and (ii) ovarian protection through treatment with gonadotropin-releasing hormone agonists (GnRHa). In France, the National Institute of Cancer (InCA) considers improving access to oncofertility services a priority (10). Efforts have been made to guarantee universal access, and all FP costs in the context of cancer are covered by the national social security system, a feature rendering French oncofertility practices unique worldwide. Nevertheless, FP counseling and access remain heterogeneous in France, as many patients receive no FP information or are not referred at the optimal time (11).

Even though it is becoming more frequent with the inflow of reassuring data (12), **pregnancy after BC** remains a rare event (13). In a recent meta-analyses, out of 711 BC survivors who received systemic treatments, 14% had a pregnancy, which is on average 40% lower than in the general population (14). However, few - if any - data is available to determine whether these low pregnancy rates are due to an actual decrease in fertility, to a lower pregnancy attempt rate due to medical reasons, or to the women’s reproductive choices. Spontaneous fertility rates after cancer treatment remain difficult to assess. The pregnancy rates achieved through the use of reproductive techniques after cancer treatments have increased with the development of FP (15–17), but recent data suggest that they remain lower after BC than in women without cancer, or women with other types of cancer (18). Furthermore, registry data show that only a few patients ask for fertility restoration (15).

The **FEERIC national study** was launched in France in March 2018 and was designed to compare fertility, pregnancy and contraception outcomes in young BC survivors and in matched cancer-free women. This online study was performed via the Seintinelles network, a collaborative social network set up to provide a source of volunteers for participation in research studies. Data were collected on twice-yearly self-completed forms. The current study focuses on BC survivors only, and is based on the enrollment’s questionnaire. The objective was to analyze patients’ attitudes and the information provided regarding fertility at the time of diagnosis, rates of referral to fertility specialists and access to FP procedures, and to describe the reproductive history between BC diagnosis and study inclusion.

## Materials and methods

### Study design

The FEERIC (Fertility, Pregnancy, Contraception after BC in France) study is a prospective study aiming at assessing the impact of BC treatment on fertility, pregnancy and contraception. The data are collected through self-administered online questionnaires via the Seintinelles* research platform (19). Seintinelles is a collaborative social network created in 2012 to accelerate the recruitment of French volunteers for cancer research studies, by connecting researchers with men and women of various ages, social and medical backgrounds, with or without a history of BC, willing to participate in research studies. The scientific board of the Seintinelles approved the FEERIC project in December 2015, and the ethics board of Sud Ouest Outre Mer II approved the project 5^th^ October, 2017.

### Patients

Patients were recruited from 12^th^ March 2018 to 27^th^ June 2019. The inclusion criteria were: female patients with a previous diagnosis of localized, relapse-free BC (invasive or *in situ*), aged 18 to 43 years at the time of diagnosis, and who had completed treatment (surgery and/or chemotherapy, and/or radiotherapy) at the time of enrollment, without any time criteria since the date of the end of treatments. Women on ongoing endocrine therapy or trastuzumab treatment were accepted. The exclusion criteria were previous hysterectomy and/or bilateral oophorectomy and/or bilateral salpingectomy. The controls were women aged from 18 to 43 years, free from BC and other cancers, who had not undergone hysterectomy and/or bilateral oophorectomy and/or bilateral salpingectomy.

### Enrollment

Cases were recruited by both (i) the Seintinelles network and (ii) eight breast care/oncofertility and gynecology centers: Institut Curie (Paris and Saint Cloud), Hôpital Saint Louis (Paris), Hôpital Cochin (Paris), Centre Léon Berard (Lyon), Centre Oscar Lambret and Centre Hospitalier Régional Universitaire (Lille), Institut Bergonie (Bordeaux). Patients were asked to participate in the study at follow-up consultations, or through the display of invitation flyers and posters in waiting rooms. In addition to the direct proposal to participate during follow-up consultations, 6 centers sent invitations to participate through mailing to patients (electronic e-mail addresses when available, or by postal mail else) (Institut Curie, Léon Bérard (Lyon), Centre Georges-François Leclerc (Dijon), Hôpital Saint-Louis (Paris), Institut du Cancer de Montpellier (Montpellier)).

### Forms

The forms were designed by workgroups including physicians involved in BC, fertility or contraceptive care, and members of the Seintinelles staff. Volunteers matching the inclusion criteria (11 questions) were sent a link to the survey and were asked to complete a baseline form at inclusion and follow-up forms every six months (a total of six forms). The baseline form contained 181 questions relating to demographic data, reproductive and contraceptive history, pregnancy plans, BC history and treatments, if applicable, data for FP procedures (FPPs) if performed, and reproductive life since the end of treatment. The questions regarding patient satisfaction were rated from 0 (unsatisfied) to 10 (extremely satisfied). In total, 28 questions specifically related to FP concerns. The database was hosted by a specialized health data provider (www.ids-assistance.com) in accordance with European General Data Protection Regulation, and was forwarded to the statistical team for analysis in a safe and anonymized manner. The current study concerns baseline characteristics and attitudes to fertility in BC survivors.

### Study endpoints

Fertility preservation procedures (FPPs) was defined as a procedure intending to help patients’s ability to have children, and included: (i) FPP with material preservation (either oocyte, embryo, or ovarian tissue cryopreservation); (ii) FPP without frozen material, *i*.*e*. GnRHa during chemotherapy; Oocyte or embryo retrieval was achieved either after controlled ovarian stimulation (COS) or without ovarian stimulation (*in vitro* maturation, IVM)). Evolutive pregnancies were defined as pregnancies evolutive after the first trimester of pregnancy. Ectopic pregnancies, miscarriage, elective abortion or abortions for medical reasons were not considered evolutive pregnancies.

For each pregnancy declared, the specific question: “was this pregnancy desired?” was asked. A pregnancy was considered desired when the patient answered “yes” and was considered unplanned if the patient answered “no”.

Median time to pregnancy was defined as time from first attempt to the occurrence of pregnancy, and median time to evolutive pregnancy was defined as time from first attempt to the occurrence of an evolutive pregnancy.

### Statistical analysis

We anticipated to include 251 patients in the FEERIC study. Thanks to a large involvement of the Seintinelles Network, a total of 517 BC patients replied to the questionnaire.

The study population was described in terms of frequencies for qualitative variables, or medians and associated ranges for quantitative variables. To compare continuous variables among different groups, Wilcoxon-Mann-Whitney test was used for groups including less than 30 patients, and for variables displaying multimodal distributions, otherwise, we used student t-test. Association between categorical variables was assessed with the chi-square test, or with the Fisher’s exact test if at least one category included less than three patients. In boxplots, lower and upper bars represent the first and third quartile, respectively, the medium bar is the median, and whiskers extend to 1.5 times the inter-quartile range. For analysis of the association between clinical variables (age, BMI, profession, study level, pregnancy plans, marital status and chemotherapy) and FPP rates, we performed a univariate analysis, with values of *p* ≤ 0.05 considered statistically significant. Analyses were performed with R software, version 3.1.2.

## Results

### Characteristics of the patients at BC diagnosis

We included 517 BC patients in the study (Table 1, figure S1). Median age at BC diagnosis was 33.6 years and median age at study inclusion was 38.0 years. In total, 455 patients (88.0%) had been educated to university level. Most patients were living with a partner at the time of diagnosis (*n*=431, 83.4% (cohabitation *n*=245, 47.4%; married *n*=186, 36.0%)). Sixty-seven patients (13.0%) were current smokers, and 368 patients (71.2%) declared no other comorbid conditions at the time of inclusion. Median BMI was 22.2 kg/m^2^ and 121 patients (23.4%) were overweight or obese. A history of infertility prior to BC diagnosis was recorded for 77 patients (14.9%) and 210 (40.6%) patients had no children at BC diagnosis. Two hundred seventy-one patients (52.4%) had a family history of BC (first degree: *n*=115 (22.2%); second or third degree: *n*=156 (30.2%)).

**Table 1:** Patient’s characteristics. Abbreviations: breast cancer (BC); body mass index (BMI); breast cancer gene (*BRCA*). Missing data: Age at BC diagnosis, n=3.

| Variable | Class | All |
| --- | --- | --- |
| *n*= |  | **517** |
| **Age at BC diagnosis** (years) |  | 33.6 [30.0, 37.1] |
| **Age at study inclusion** (years) |  | 38.0 [34.0, 41.0] |
| **Age** (by class) | <30 | 38 (7.4) |
|  | [30-35) | 132 (25.5) |
|  | [35-40) | 215 (41.6) |
|  | >=40 | 132 (25.5) |
| **Study level** | High school or lower | 62 (12.0) |
|  | University | 455 (88.0) |
| **Profession** (class) | Intermediate | 221 (42.7) |
|  | Low or unemployed | 48 (9.3) |
|  | Superior | 248 (48.0) |
| **Marital status** (at BC diagnosis) | Couple | 431 (83.4) |
|  | *Relationship* |  |
|  | Cohabitation | 245 (56.8) |
|  | Married | 186 (43.2) |
|  | Single | 86 (16.6) |
| **BMI** (continuous) |  | 22.2 [20.4, 24.6] |
| **BMI** (4 classes) | Underweight | 30 (5.8) |
|  | Normal weight | 366 (70.8) |
|  | Pre-obesity | 84 (16.2) |
|  | Obesity | 37 (7.2) |
| **Smoking status** | Current | 67 (13.0) |
|  | Former | 207 (40.0) |
|  | Never | 243 (47.0) |
| **Comorbidity** | No | 368 (71.2) |
|  | Yes | 149 (28.8) |
| **Co-medications** | No | 401 (77.6) |
|  | Yes | 116 (22.4) |
| **Infertility history** | No | 440 (85.1) |
|  | Yes | 77 (14.9) |
| **Previous pregnancy** | No | 150 (29.0) |
|  | Yes | 367 (71.0) |
| **Number of children** (3 classes) | 0 | 210 (40.6) |
|  | 1 | 119 (23.0) |
|  | More than 1 | 188 (36.4) |
| **Previous births** | No | 210 (40.6) |
|  | Yes | 307 (59.4) |
|  | *Breastfeeding history* |  |
|  | No | 81 (26.4) |
|  | Yes | 226 (73.6) |
|  | *Breastfeeding duration* (months) | 4.0 [2.25, 9.00] |
| **Familial history of BC** | No | 246 (47.6) |
|  | At least 1 first-degree relative | 115 (22.2) |
|  | At least 1 second-degree relative | 156 (30.2) |
| **Oncogenetic counseling** | No | 98 (19.0) |
|  | Yes | 419 (81.0) |
| **Genetic testing** | No | 107 (20.7) |
|  | Yes | 410 (79.3) |
|  | ***Oncogenetic results*** |  |
|  | No mutation | 287 (70.0) |
|  | BRCA1 | 39 (9.5) |
|  | BRCA2 | 22 (5.4) |
|  | Others | 4 (1.0) |
|  | Results awaited / not known | 58 (14.1) |

Most of the patients (*n*=419, 81.0%) had received oncogenetic counseling and 410 (79.3%) had undergone genetic testing. No constitutional mutation was found in 287 (70.0%) (oncogenetic results available at the time of study inclusion, n=352), whereas germline mutations were detected in 61 patients (*BRCA1 n*=39 (9.5%); *BRCA2* mutation *n*=22 (5.4%); others *n*=4 (1.0%)).

### BC treatments

Median time from BC diagnosis to inclusion was 34.9 months (Supplemental table 1). The first-line treatment was surgery in 66.3% of the patients (*n*=343), whereas 33.5% underwent first-line systemic treatment (*n*=173) (Fig. 1A). All but one of the patients underwent surgery for BC. Surgical procedures were mastectomy n=239 (46.5%) / lumpectomy n=275 (53.5%), axillary node dissection n=252 (54.2%) / sentinel node biopsy n=213 (45.8%). Chemotherapy was administered to 424 patients (82.0%), mostly with anthracyclines followed by taxanes regimens, 172 (40.6%) in a neoadjuvant setting (including 9 with adjuvant treatment, 2.2%) and 252 (59.4%) in an adjuvant setting. In total, 443 (85.7%) patients received radiotherapy, 126 (24.4%) patients received trastuzumab treatment for *HER2*-positive cancers, and 330 (63.8%) received endocrine therapy (Fig. 1A). Figure 1B displays the different care pathways for the entire cohort.

**Figure 1:**
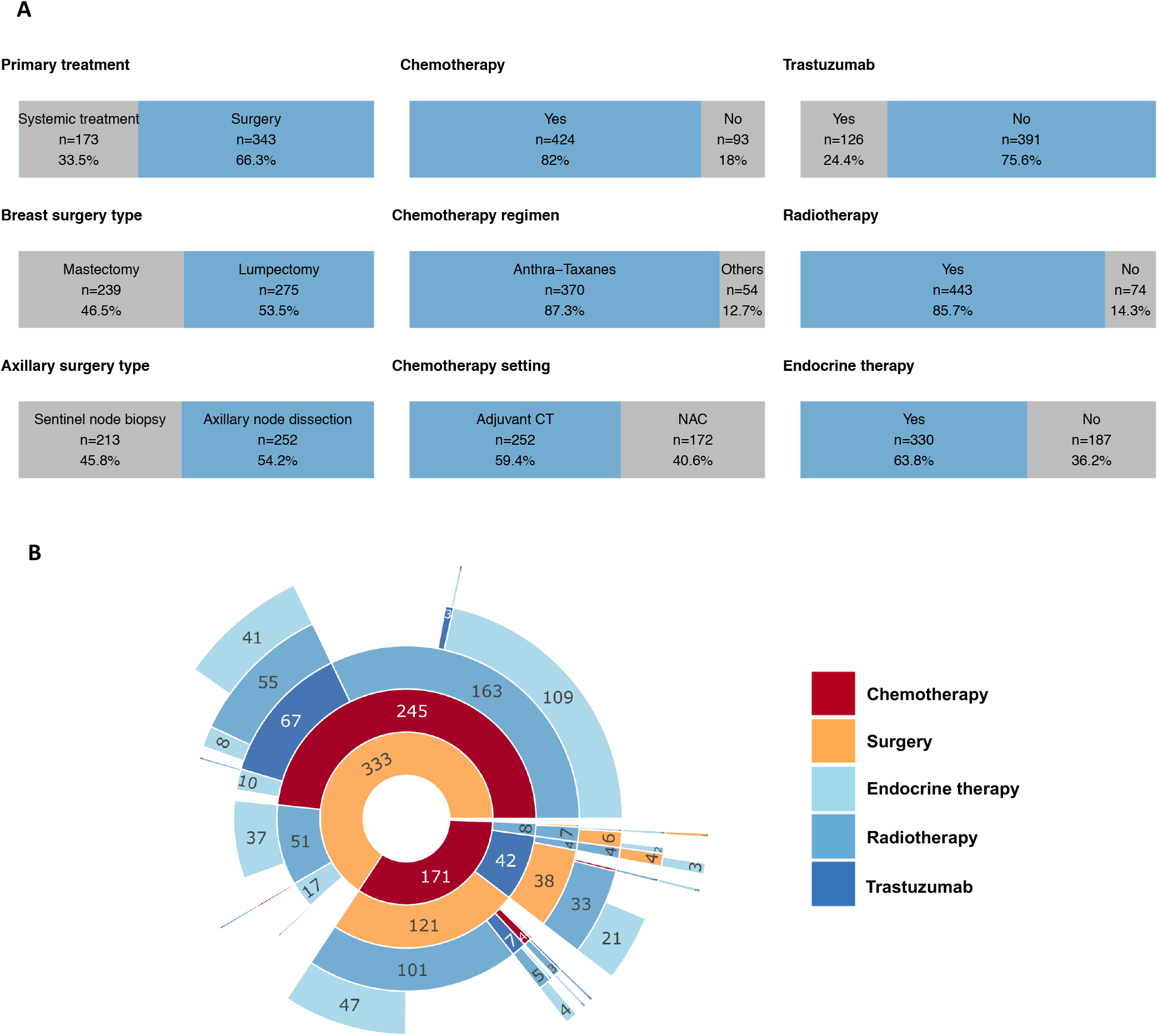
Breast cancer treatments and care pathway. A, Repartition of BC treatments. B, Chart representing the sequential care pathways. Twelve patients were discarded from the sunburst because of discrepancies between the treatment dates mentioned in the study and the pathway. Abbreviations: human epidermal growth factor receptor 2 (Her2), neoadjuvant chemotherapy (NAC).

### Pregnancy plans, fertility counseling and oncofertility preservation procedures at BC diagnosis

At the time of BC diagnosis, 218 patients (42.2%) declared no wish of pregnancy, 206 stated they could consider a pregnancy in the future (39.8%), and 67 (13.0%) reported that they were trying to become pregnant (Fig. 2A, supplemental table 2). BC was diagnosed during pregnancy in 26 patients (5.0%), and the outcome of the pregnancy was favorable in most cases (full-term pregnancy, *n*=17 (65.4%); elective abortion, *n*=3 (11.5%); abortion for medical reasons, *n*=3 (11.5%); miscarriage, *n*=3 (11.5%)) (supplemental table 2).

**Figure 2:**
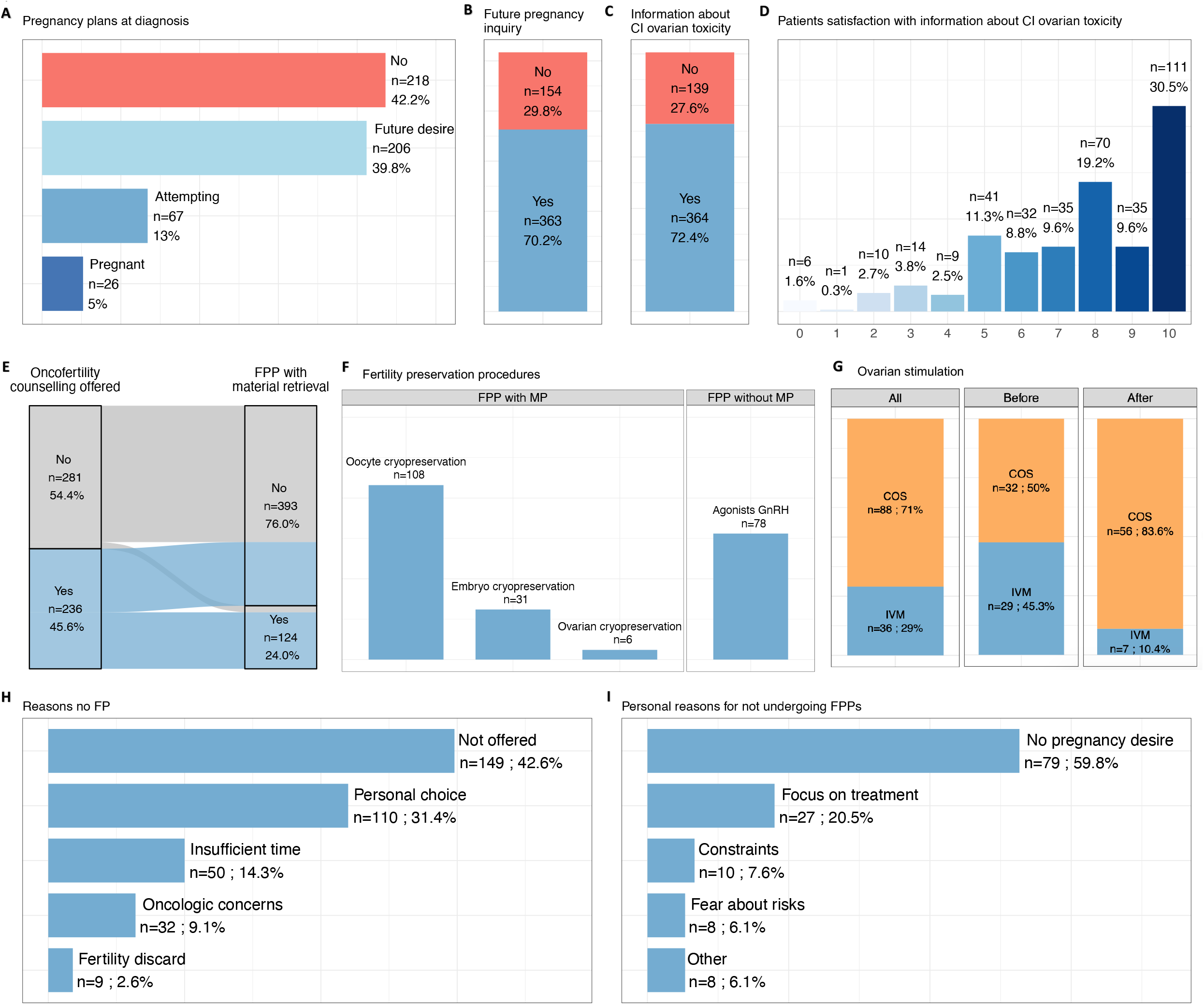
Oncofertility counseling, fertility preservation procedures and decision making. A, Repartition of patients’ pregnancy plans at diagnosis. B, Barplot of medical inquiry about patient’s future pregnancy desire. C, Barplot representing the proportion of women who received an information about chemotherapy-induced (CI) ovarian toxicity. D, Patient’s satisfaction towards information on chemotherapy-induced (CI) ovarian toxicity (0-to-10 Numerical Rating Scale (NRS), where 0 is strongly dissatisfied and 10 is fully satisfied), out of 364 patients who received information. E, Sankey diagram representing the association between oncofertility counseling proposal and completion of a fertility preservation procedure (FPP). Thirteen patients had a FPP with material preservation by their own initiative in the absence of counselling being offered by oncology team. F, Repartition of FPP with or without material preservation (MP). A combination of methods was used in 39 patients: mostly oocyte cryopreservation plus GnRHa treatment (*n*=15), followed by the cryopreservation of oocytes and embryos (*n*=13). G, Barplots representing ovarian stimulation performance according to chemotherapy setting. H, Repartition of the reasons for not beneficiating from a FPP. Main reason was the absence of offer (n=149; 42.6%). I, Repartition of the personal reasons for not undergoing a FPP. Abbreviations: chemotherapy-induced (CI); gonadotropin-releasing hormone agonists (GnRHa); controlled ovarian stimulation (COS); in vitro maturation (IVM); neoadjuvant chemotherapy (NAC); chemotherapy (CT); fertility preservation (FP); fertility preservation procedure (FPP); material preservation (MP).

A health care practitioner inquired if the patient would consider future pregnancy after treatment in 363 patients (70.2%) (Fig. 2B) and 72.4% (n=364) patients recalled being informed about chemotherapy-induced ovarian toxicity (Fig. 2C). Among them, the degree of satisfaction of this information was high (median score: 8/10) (Fig. 2D). Specialized oncofertility counseling was offered to 236 patients (45.6%) and 124 women (24.0%) underwent at least one FPP with material preservation (Fig. 2E). The distribution of procedures was as follows: oocyte cryopreservation (*n*=108, 87.1%), embryo cryopreservation (*n*=31, 25.0%), ovarian cryopreservation (*n*=6, 4.8%) (Fig. 2F). In addition, GnRHa were used during treatment in 78 patients (15.1%), representing a total of 181 patients with at least one FPP performed (35.0%) (Fig. 2F). Eighty-eight patients (71.0%) had material preservation after controlled ovarian stimulation, while 36 patients had an *in vitro* maturation procedure (29.0%) (supplemental table 2). Controlled ovarian stimulation was less frequent for FPPs performed in the before surgery (*n*=30, 37.5%) than for FPPs in after surgery (*n*=50, 62.5%) (Fig. 2G).

Age and year of BC diagnosis (*p*<0.001), study level (*p*=0.003), previous children (*p*<0.001), plans for pregnancy at diagnosis (*p*<0.001) and BC treatment with chemotherapy (*p*=0.001) were significantly associated with the likelihood of undergoing a FPP procedure with material preservation (supplemental table 3). The reasons for which women did not undergo FPP procedures based on cryopreservation techniques was a lack of such procedures being offered (*n*=149, 42.6%), personal choice of the patient (*n*=110, 31.4%), insufficient time available (*n*=50, 14.3%), oncological concerns (*n*=32, 9.1%), and exclusion on the base of fertility conditions (*n*=9, 2.6%) (Fig. 2H). The personal reasons for not undergoing FPPs cited by patients included not wanting to have a baby (*n*=79, 59.8%), followed by a desire to focus on BC treatment (*n*=27, 20.5%), constraints (*n*=10, 7.6%) or fears (*n*=8, 6.1%) relating to treatment (Fig. 2I).

### Gynecological and reproductive history since BC treatment

#### Amenorrhea, endocrine therapy, couple situation

Of the 424 patients who received CT, 406 (95.8%) reported amenorrhea during treatment (median duration: 11 months), and the duration of amenorrhea increased with age (Fig. 3A; supplemental table 4). Amenorrhea was reversible in most patients (*n*=317, 74.8%) (supplemental table 4). Endocrine treatment was stopped prematurely (median: 28 months) in 99 of 330 patients (30.0%), either temporarily (*n*=77, 23.3%) or definitively (*n*=22, 6.7%). The main reasons for stopping endocrine treatment were a desire to become pregnant (*n*=51, 51.5%), adverse side effects (*n*=35, 35.4%), or other reasons (*n*=16, 16.2%). Median time of endocrine therapy intake was significantly shorter in patients who stopped due to side effects than in patients who stopped because of a pregnancy desire (19.8 months *versus* 36.4 months respectively, *p*<0.001)

**Figure 3:**
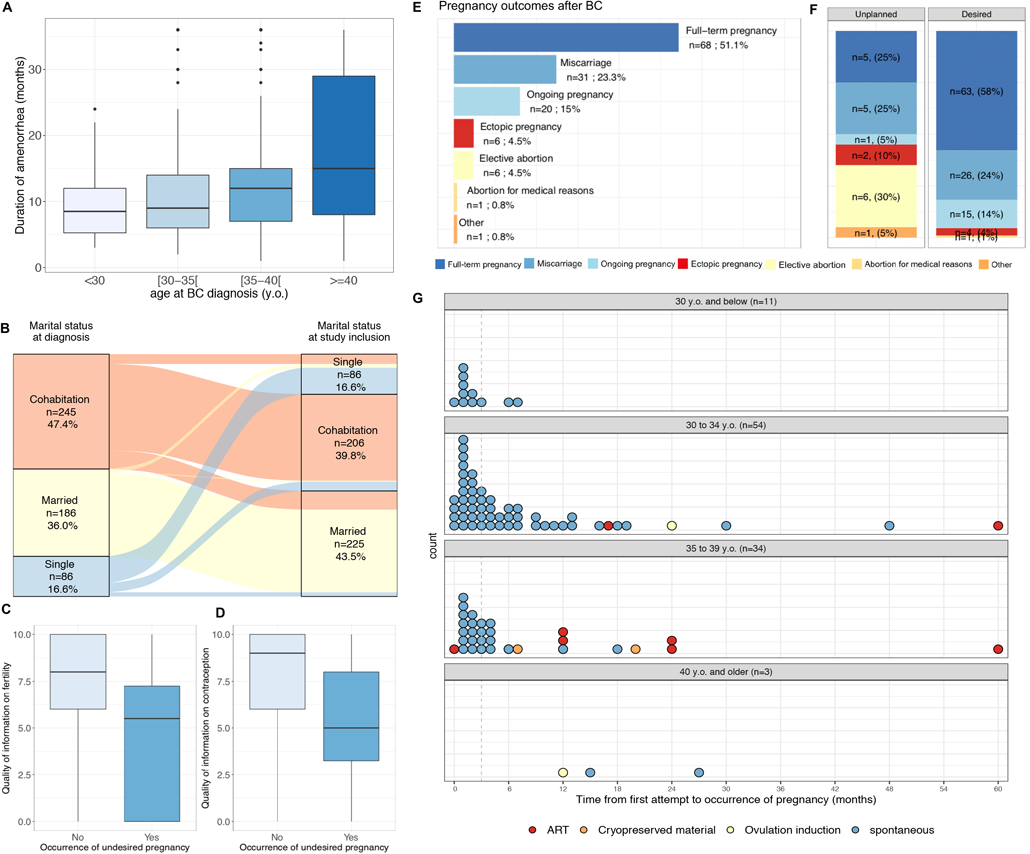
Occurrence and outcome of pregnancies after BC treatment. A, Association between age and duration of amenorrhea. Median duration of amenorrhea is as follows: <30 y.o.: 8 months; [30-35[ :9 months; [35-40[: 12 months; >=40: 15 months. B, Sankey diagram of the evolution of marital status between diagnosis and study inclusion. C, Association between the information received on fertility and the occurrence of an unplanned pregnancy. D, Association between the information received on contraception and the occurrence of an unplanned pregnancy. E, Outcomes of the 133 pregnancies achieved after breast cancer in the FEERIC cohort. F, Barplots of pregnancy outcomes depending on the status unplanned (n=20) or desired (n=109) and the pregnancy occurrence (spontaneous n=113 or use of assisted reproductive technology n=16) out of the 129 pregnancies for which the unplanned/desired status was known. G, Time from first attempt to pregnancy occurrence in months according to age classes and pregnancy achievement method out of the 102 patients for which the attempt duration was known. Abbreviations: years old (y.o.); assisted reproductive technology (ART); breast cancer (BC)

Between the BC diagnosis and the inclusion of the study, 19.3% of patients (n=100) changed marital status (Fig. 3B). The proportion of patients who formed new couples (29/86; 33.7%) was higher than the proportion of patients who were initially in a couple but became single (29/402; 7.2%) (Fig. 3B).

#### Unplanned pregnancies

During a median follow-up of 26.7 months between the end of BC treatment and inclusion in the study, 18 patients (3.5%) experienced an unplanned pregnancy. Patients who had an unplanned pregnancy were significantly younger at BC diagnosis (*p*=0.03), were more likely to have previous pregnancies (*p*=0.01) or previous children at BC diagnosis (*p*=0.02), were less likely to have received fertility counseling (*p*=0.04) and tended to have less contraceptive counseling (*p*=0.08) (Supp table 5). The satisfaction regarding information on fertility and contraception was significantly lower in patients who had subsequent unplanned pregnancy than in patients who did not (*p*=0.021 and *p*<0.001 respectively) (supplemental table 5).

#### Pregnancies after BC

Overall a total of 133 pregnancies occurred in 85 patients (16.4%), with 20 of them (15.0%) being unplanned). Pregnancy onset was spontaneous in 113 cases (87.6%), and medical intervention was required for 16 pregnancies (ART *n*=10 (13.8%); oocyte donation *n*=2; use of cryopreserved material obtained after controlled ovarian stimulation *n*=2; and ovulation induction *n*=2) (supplemental table 6). One of the two pregnancies achieved with cryopreserved material ended in a live birth; the other pregnancy was still underway at the time of inclusion in the study. The outcomes of pregnancies were as follows: full-term pregnancy, *n*=68 (51.1%); miscarriage, *n*=31 (23.3%); ongoing pregnancy, *n*=20 (15.0%) ectopic pregnancy, *n*=6 (4.5%); elective abortion, *n*=6 (4.5%); abortion for medical reasons, *n*=1 (0.8%), other *n*=1 (0.8%) (Fig. 3E).

The outcome of the pregnancy was significantly different according to whether the pregnancy was unplanned or desired (*p*<0.001), with notably a higher proportion of elective abortions in unplanned pregnancies than in desired pregnancies (30.0% *versus* 0% respectively) (Fig. 3F). There were very few significant obstetrical complications (twins *n*=2, gestational diabetes *n*=3, pre-eclampsia *n*=2, preterm birth *n*=2, risk of preterm delivery *n*=1, post-partum peritonitis *n*=1, CMV seroconversion *n*=1) and no malformations was reported in the infants. The infants were delivered by the vaginal route (*n*=55; 80.9%) or by cesarean section (n=13; 19.1%) (supplemental table 6).

#### Pregnancies attempts after BC and time to pregnancy

Overall, since the end of treatment, 127 patients (24.6%) had attempted to get pregnant; 78 of these patients (61.4%) had at least one pregnancy, 52 patients had at least one live birth (40.9%) and 20 (25.6%) were pregnant at inclusion in the study (five patients were pregnant on inclusion and also had one prior live birth during the follow-up period). For the 109 desired pregnancies, both median time to pregnancy (TTP) and median time to evolutive pregnancy were three months, and 68.6% of the patients had conceived in the 6 months following first attempt. Median TTP was significantly higher in patients aged 40 and older (median TTP: 15 months), than in younger women (median TTP <30 y.o.: 1 month; 30 to 34 y.o.: 3.5 months; 35 to 39: 3 months, *p*=0.02) (Fig. 3G). At study inclusion, 66 BC patients (12.8%) declared that they were still currently trying to get pregnant.

## Discussion

This large study on young BC survivors provides important insights into patients’ attitudes and expectations towards fertility and pregnancy, as well as oncofertility practices nationwide in France.

***First***, one third of women declared that they had not been informed of possible fertility damage before treatment, nor were they asked about future pregnancies plans after BC treatment. Academic societies strongly recommend informing patients about oncofertility (10,16,17), but little was known to date about the prevalence of counseling in French BC survivors. American data have shown that 47 to 68% receive information about FP (18–21). In France, one single-center retrospective study showed that only four of 230 (1.7%) BC patients treated between 2000 and 2010 received information before treatment, with another 49 (21.7%) receiving information after gonadotoxic treatments had been initiated (22). Another French national survey (23) on 104 female cancer patients under the age of 45 years diagnosed in 2002 reported that 31 patients (30.0%) had not been informed about the risk of infertility before treatment. Finally, in the VICAN study (24), which included 427 women aged from 18 to 40 years treated in 2010, 291 women (68.1%) reported having received no FP counseling before cancer treatment.

In line with the lack of information on chemotherapy-induced ovarian damage, almost one third of the patients of our cohort reported not undergoing FPPs because this option was not offered (*n=*149, 28.8%), or because too little time was available (*n*=50, 9.7%). Both these obstacles to FPPs are avoidable and represent actionable ways of improving patient care. On the other hand, a significant subset of women (*n*=110, 21.3%) decided not to go through with FPPs despite receiving counseling. FPPs are costly and the low rates of referral in other studies have been linked to the financial burden (26). However, this bias can be ruled out here, because all French citizens are covered by a universal social security system guaranteeing the full reimbursement of FP fees.

***Second***, the main technique used was oocyte cryopreservation (87.1%), which was much more frequently used than embryo (25.0%) or cortex cryopreservation (4.8%). This distribution seems to be clinically relevant, as, in addition to yielding similar results to embryo freezing for efficacy (9), oocyte cryopreservation has the key advantage of preserving the fertility of the woman, rather than the fertility of the couple (3). Recent studies have not found divorce or separation rates to be particularly high following BC diagnosis (27,28), nevertheless the preservation of the woman’s fertility (oocyte), rather than that of the couple (embryo), is particularly advantageous for single women or women whose relationships are breaking down. Surprisingly, our results showed that nearly half of patients who had FPP with material preservation in the neoadjuvant setting had ovarian stimulation. This result was unexpected, as ovarian stimulation (OS) is not recommended before breast tumor excision in France (3), and there is only limited evidence concerning the safety of OS in the neoadjuvant setting (20,21). Despite the disappointingly low birth rates previously reported after IVM (12,30), approximatively 30% of the patients had IVM. As this rate is expected to increase due to the widespread use of neoadjuvant chemotherapy in young women, results of such techniques should be monitored closely to assess the efficacy of this procedure with long term follow-up. Of note, only 15.1% of patients received GnRHa treatment together with chemotherapy. This low rate may be accounted for by the study period during which most of the patients were treated, as only 117 patients from the cohort (22.6%) were treated in or after 2017. Indeed, the pre-2017 guidelines considered GnRHa to be an experimental technique (31–33), and GnRHa has been considered a valid option for FP in the National Comprehensive Cancer Network (NCCN) and ASCO guidelines only since 2017 and 2018, respectively (34). Two randomized controlled trials (35,36) have since reported significantly lower rates of post-chemotherapy ovarian failure in patients receiving GnRHa during chemotherapy, for both hormone receptor-positive and - negative BC, leading to an update of NCCN guidelines in 2019 (37). All the patients of our study were treated before 2019, and presumably, this treatment will be increasingly used in routine care as an ovary- protecting agent, in addition to other fertility preservation methods.

***Third***, with a rather short median time between the end of treatment and inclusion, 16.4% of the patients of the cohort had already experienced at least one pregnancy, most of which were spontaneous, and only 2 pregnancies occurred after use of cryopreserved material. The occurrence of unwanted pregnancies (3.9%) was even more frequent than the use of ART (3.1%). These results highlight the statement that contraception is as important as FP in young breast cancer patients (38). In addition, the quality of the fertility and the contraceptive counseling was significantly associated with a lower likelihood of experiencing unplanned pregnancies. Properly addressing this topic could spare women the burden of undergoing elective abortions or abortions for medical reasons. Finally, for patients who became pregnant after BC, the median time between pregnancy desire and its occurrence was short (three months). Although we cannot formally exclude the possibility that a subset of women currently attempting to get pregnant may remain genuinely infertile due to chemotherapy-induced ovarian damage, the data provided here are reassuring concerning the likelihood of spontaneous pregnancies after BC.

Our study has several strengths. It reports one of the largest recent study on fertility concerns and attitudes at BC diagnosis based on patient reported outcomes. It also provides unprecedented data on unplanned pregnancies, and time between pregnancy attempt and pregnancy occurrence, and provides relative comparison in the number of pregnancies occurring after PF material reuse *versus* spontaneously in a real-world cohort. It also has limitations. Our data could have overestimated the information rate given in the whole young breast cancers population, as most of the FEERIC study women came from high-level socio-professional backgrounds. A similar bias has been reported in other studies recruiting patients via online networks (18,25). Moreover, study inclusions performed by cancer centers or teaching hospitals might not exactly reflect the field reality. Finally, responders to our study may represent a subset of BC patients with a particular interest in fertility following BC, and underrepresent either patients who experienced relapse or those who already have completed their parental project.

***In conclusion***, our work provides an overview of the fertility and pregnancy concerns in a selected cohort of young French BC survivors. Information about and access to FPPs could be further improved. Healthcare professionals should be aware of the need to systematically offer fertility counseling to patients before BC treatment. In addition, care pathways should be optimized to facilitate rapid access to FPPs if desired by the patient and feasible. Finally, contraception should also be considered a critical topic at BC diagnosis and during the follow-up, as unplanned pregnancies represent a preventable burden for BC survivors. Finally, the longitudinal follow-up of the FEERIC cohort for three years will make it possible to determine whether pregnancy rates differ between BC survivors and matched controls.

## Data Availability

Data are available on request due to privacy

## Acknowledgments

The FEERIC study was funded by Institut du Cancer InCA, InCA-SHS, grant No. 2016-124. The authors thank all the study participants from the Seintinelles Network, and Lili Sohn who is the sponsor of the study.

## Figure titles and abbreviations (intended for color reproduction on the Web and in print)

<u>Supplemental figure 1: Patient characteristics of the 517 patients at the time of BC diagnosis.</u> A, Median age at BC diagnosis was 33.6 years. B, Median age at study inclusion was 38.0 years. C, Socio-economic level at diagnosis. D, Marital status at diagnosis. E, Smoking status at diagnosis. F, Presence of comorbidities at diagnosis. G, BMI at diagnosis, median was 22.2 kg/m^2^. H, Number of children at diagnosis. I, Family history of BC. J, Presence of a genetic mutation among 410 patients who had oncogenetic screening. Abbreviations: breast cancer (BC); body mass index (BMI); Breast Cancer gene (*BRCA*)

## Table titles and abbreviations

**Supplemental table 1:** BC treatments. All but one patient had surgery. Abbreviations: breast cancer (BC); human epidermal growth factor receptor 2 (Her2). Missing data: Time from BC diagnosis to inclusion, n=3; BC surgery, n=3; Axillar surgery, n=52

| Variable | Class | All |
| --- | --- | --- |
| *n*= |  | **517** |
| **Time from BC diagnosis to inclusion (months)** |  | 34.9 [18.7, 64.1] |
| **Primary treatment** | Surgery | 343 (66.3) |
|  | Neoadjuvant treatment | 173 (33.5) |
|  | Chemotherapy | 172 (33.3) |
|  | Trastuzumab | 43 (8.3) |
|  | Endocrine therapy | 2 (0.4) |
|  | Radiotherapy | 12 (2.3) |
| **BC surgery (type)** | Lumpectomy | 275 (53.5) |
|  | Mastectomy | 239 (46.5) |
| **Axillary surgery (type)** | Axillary node dissection | 252 (54.2) |
|  | Sentinel node biopsy | 213 (45.8) |
| **Chemotherapy** | No | 93 (18.0) |
|  | Yes | 424 (82.0) |
|  | **Chemotherapy regimen** |  |
|  | Anthra-taxans | 370 (87.3) |
|  | Others | 54 (12.7) |
|  | **Chemotherapy setting** |  |
|  | NAC | 163 (38.4) |
|  | NAC & adjuvant CT | 9 (2.1) |
|  | Adjuvant CT | 252 (59.4) |
| **Radiotherapy** | No | 74 (14.3) |
|  | Yes | 443 (85.7) |
| **Anti-HER2 therapy** | No | 391 (75.6) |
|  | Yes | 126 (24.4) |
| **Endocrine therapy** | No | 187 (36.2) |
|  | Yes | 330 (63.8) |
|  | *At least one intake* | 317 (96.1) |

**Supplemental table 2:** Fertility status, oncofertility counseling and procedures at BC diagnosis. Nine patients underwent FPPs despite not having chemotherapy. Abbreviations: gonadotropin-releasing hormone agonists (GnRHa); chemotherapy (CT); fertility preservation procedure (FPP); *in vitro* maturation (IVM)

| Variable | Class | All |
| --- | --- | --- |
| **n=** |  | **517** |
| **Pregnancy plans at BC diagnosis** | Attempting pregnancy | 67 (13.0) |
|  | Future pregnancy desire | 206 (39.8) |
|  | No | 218 (42.2) |
|  | Pregnant at diagnosis | 26 (5.0) |
|  | ***Outcomes of pregnancy*** |  |
|  | Full-term pregnancy | 17 (65.4) |
|  | Elective abortion | 3 (11.5) |
|  | Abortion for medical reasons | 3 (11.5) |
|  | Miscarriage | 3 (11.5) |
| **Future pregnancy inquiry**<br>(by any health care provider) | No | 154 (29.8) |
|  | Yes | 363 (70.2) |
| **Information about chemotherapy induced ovarian toxicity** *out of 503* | No | 139 (27.6) |
|  | Yes | 364 (72.4) |
| **Quality of information** (from 1 to 10)<br>(*median*) |  | 8 [6 - 10] |
| **Oncofertility counselling offered** | No | 281 (54.4) |
|  | Yes | 236 (45.6) |
| **Oncofertility counselling performed** | No | 312 (60.3) |
|  | Yes | 205 (39.7) |
| **FPP with material preservation** | Material preserved | 124 (24.0) |
|  | *Oocyte cryopreservation* | 108 |
|  | *Embryo cryopreservation* | 31 |
|  | *Ovarian cortex cryopreservation* | 6 |
|  | Ovarian stimulation |  |
|  | No (IVM) | 36 (29.0) |
|  | Yes | 88 (71.0) |
| **FPP without material preservation** | LHRH agonists during CT | 78 (15.1) |
| **Any FPP** | No | 336 (65.0) |
|  | Yes | 181 (35.0) |
| **FPP timing** | Before surgery | 64 (48.9) |
|  | After surgery | 67 (51.1) |

**Supplemental table 3:** Association between clinical characteristics and the performance of fertility preservation procedures. Abbreviations: breast cancer (BC); body mass index (BMI); breast cancer gene (*BRCA*). Missing data: year of BC diagnosis, n=3

| Variable | Class | No | Yes | *p* |
| --- | --- | --- | --- | --- |
| *n*= 517 |  | 393 | 124 |  |
| **Age at BC diagnosis** |  | 34.0 (4.7) | 31.8 (3.9) | **<0.001** |
| **Age at BC diagnosis**<br>(class) | <30 | 19 (4.8) | 19 (15.3) | **<0.001** |
|  | [30-35[ | 80 (20.4) | 52 (41.9) |  |
|  | [35-40[ | 170 (43.3) | 45 (36.3) |  |
|  | >=40 | 124 (31.6) | 8 (6.5) |  |
| **Year BC diagnosis** | <=2010 | 50 (12.8) | 1 (0.8) | **<0.001** |
|  | 2010-2015 | 195 (50.0) | 60 (48.4) |  |
|  | >2015 | 145 (37.2) | 63 (50.8) |  |
| **Study level** | Bachelor or lower | 57 (14.5) | 5 (4.0) | **0.003** |
|  | University | 336 (85.5) | 119 (96.0) |  |
| **Profession** (class) | Intermediate | 178 (45.3) | 43 (34.7) | 0.105 |
|  | Low CSP or unemployed | 36 (9.2) | 12 (9.7) |  |
|  | Superior | 179 (45.5) | 69 (55.6) |  |
| **BMI** (4 classes) | Underweight | 23 (5.9) | 7 (5.6) | 0.902 |
|  | Normal weight | 275 (70.0) | 91 (73.4) |  |
|  | Pre-obesity | 66 (16.8) | 18 (14.5) |  |
|  | Obesity | 29 (7.4) | 8 (6.5) |  |
| **Smoking status** | No | 341 (86.8) | 109 (87.9) | 0.861 |
|  | Yes | 52 (13.2) | 15 (12.1) |  |
| **BRCA mutation genes** |  | 43 (8.3) | 22 (4.3) | 0.358 |
|  | BRCA1 | 24 (55.8) | 15 (68.2) |  |
|  | BRCA2 | 17 (39.5) | 5 (22.7) |  |
|  | Others | 2 (4.7) | 2 (9.1) |  |
| **Infertility history** | No | 334 (85.0) | 106 (85.5) | 0.99 |
|  | Yes | 59 (15.0) | 18 (14.5) |  |
| **Previous children** | No | 120 (30.5) | 90 (72.6) | **<0.001** |
|  | Yes | 273 (69.5) | 34 (27.4) |  |
| **Marital status** (BC diagnosis) | Single | 60 (15.3) | 26 (21.0) | 0.178 |
|  | Coupled up | 333 (84.7) | 98 (79.0) |  |
| **Attitude towards pregnancy desire** | Attempted pregnancy | 44 (11.2) | 23 (18.5) | **<0.001** |
|  | Future pregnancy desire | 128 (32.6) | 78 (62.9) |  |
|  | No | 196 (49.9) | 22 (17.7) |  |
|  | Pregnant at diagnosis | 25 (6.4) | 1 (0.8) |  |
| **Chemotherapy** | No | 84 (21.4) | 9 (7.3) | **0.001** |
|  | Yes | 309 (78.6) | 115 (92.7) |  |

**Supplemental table 4:** Amenorrhea, endocrine therapy, and marital status at study inclusion. Abbreviations: breast cancer (BC); chemotherapy (CT)

| Variable | Class | All |
| --- | --- | --- |
| *n*= |  | **517** |
| **Time from end of treatment to inclusion**<br>(months) |  | 35.8 (32.4) |
| **Amenorrhea during CT** |  | 424 (82.0) |
|  | No | 18 (4.2) |
|  | Yes | 406 (95.8) |
|  | *Amenorrhea duration (months)* | 11.0 [6.0, 18.0] |
|  | *Amenorrhea reversibility* |  |
|  | No | 107 (25.2) |
|  | Yes | 317 (74.8) |
| **Endocrine therapy early discontinuation** | Yes | 99 (30.0) |
|  | *Definitively* | 22 (6.9) |
|  | *Temporarily* | 77 (24.3) |
| **Marital status** (current) | Cohabitation | 206 (39.8) |
|  | Married | 225 (43.5) |
|  | Single | 86 (16.6) |

**Supplemental table 5:**
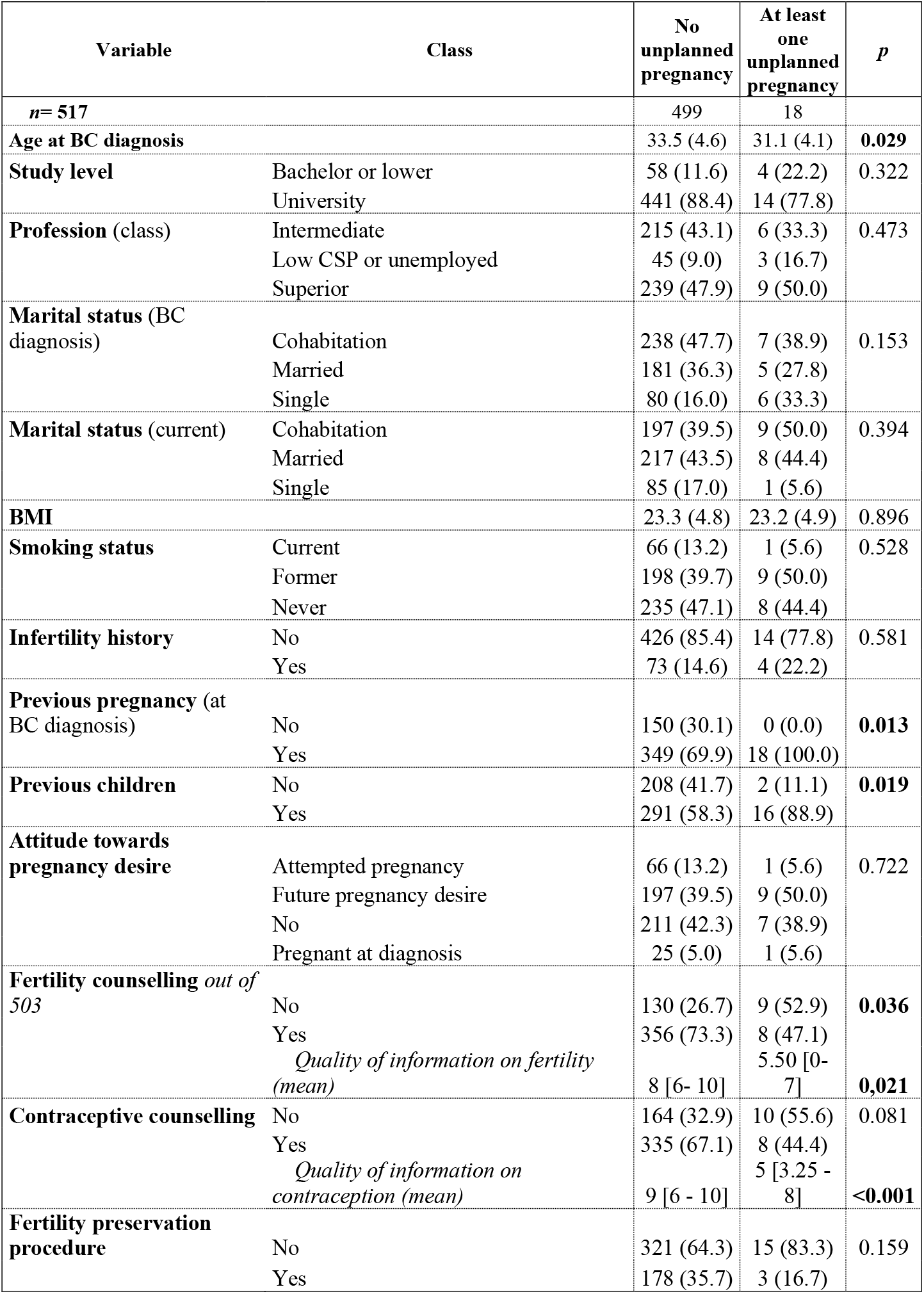
Factors associated with the occurrence of an unplanned pregnancy. Abbreviations: breast cancer (BC); body mass index (BMI)

**Supplemental table 6:** Occurrence and outcome of pregnancies after BC treatment. Abbreviations: assisted reproductive technology (ART). Missing data: Pregnancy planning, n=4, Pregnancy onset, n=4

| Variable | Class | All |
| --- | --- | --- |
| **Pregnancies** (in 85 patients) |  | **133** |
| **Pregnancy planning** *out of 129* | Unplanned (in 18 patients) | 20 (15.5) |
|  | Desired | 109 (84.5) |
| **Pregnancy onset** *out of 129* | Spontaneous | 113 (87.6) |
|  | Medical intervention | 16 (12.4) |
|  | *ART* | 10 (62.5) |
|  | *Oocyte donation* | 2 (12.5) |
|  | *Cryopreserved material reuse* | 2 (12.5) |
|  | *Ovulation induction* | 2 (12.5) |
| **Pregnancy outcome** | Ectopic pregnancy | 6 (4.5) |
|  | Induced abortion | 6 (4.5) |
|  | Medical abortion | 1 (0.8) |
|  | Miscarriage | 31 (23.3) |
|  | Ongoing pregnancy | 20 (15.0) |
|  | Other | 1 (0.8) |
|  | Full-term pregnancy | 68 (51.1) |
|  | ***Delivery Route*** |  |
|  | Caesarean section | 13 (19.1) |
|  | Vaginal birth | 55 (80.9) |
| **Obstetrical conditions** *out of 12* | Twins | 2 (1.6) |
|  | Pre-eclampsia | 2 (1.6) |
|  | Preterm birth | 2 (1.6) |
|  | Gestational diabetes | 3 (2.4) |
|  | Risk of preterm birth | 1 (0.8) |
|  | Post-partum peritonitis | 1 (0.8) |
|  | CMV seroconversion | 1 (0.8) |

