## Supplementary figures and images for "Pregnancy, fertility concerns, and fertility preservation procedures in French breast cancer survivors in the FEERIC national study (on behalf of the Seintinelles research network)"

### Supplemental table 1

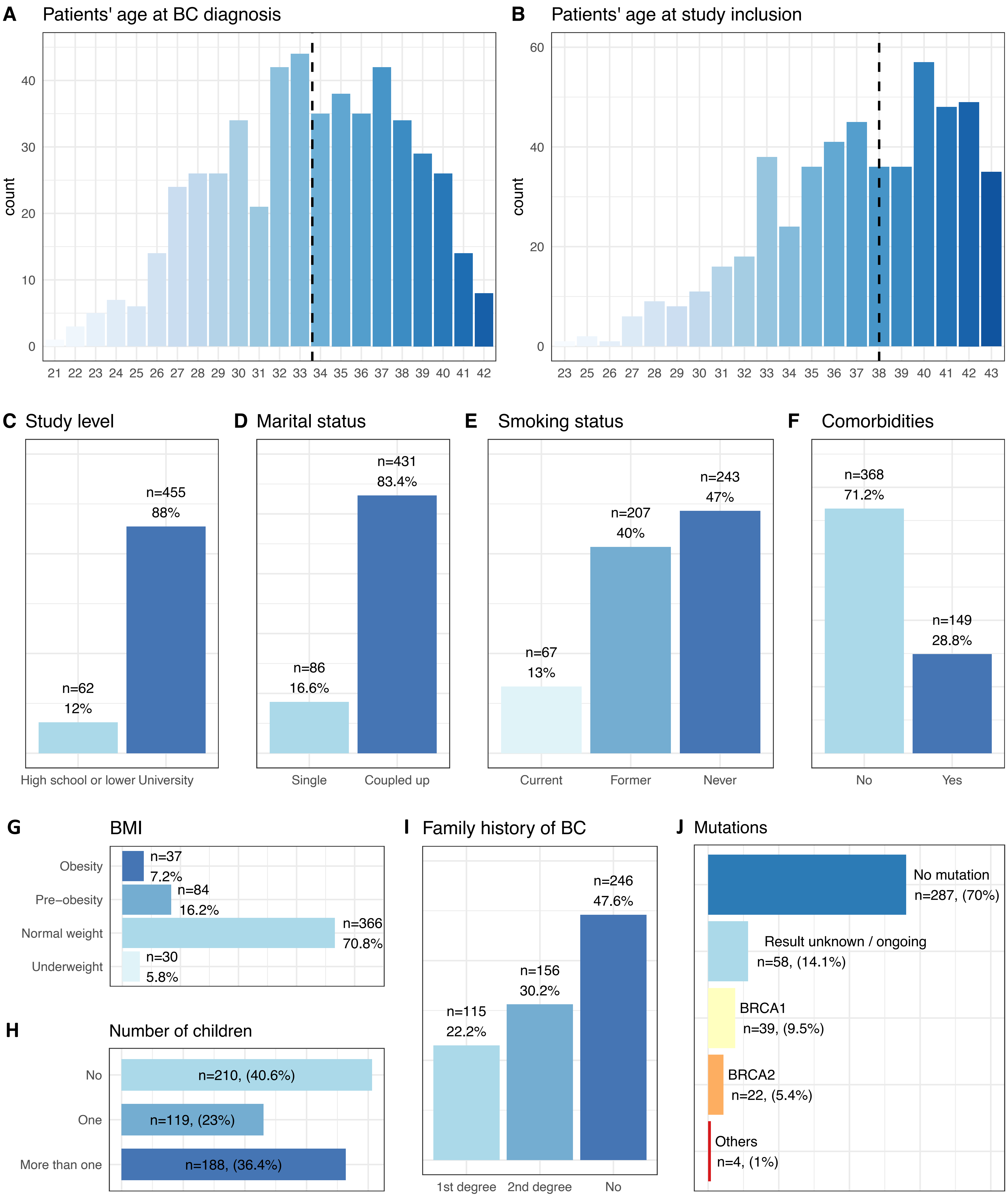
